# Automated Net Water Uptake Quantification in Ischemic Stroke: Validation Against Manual Measurement in the AcT Trial

**DOI:** 10.64898/2026.07.08.26357599

**Authors:** Sanraj Singh, Pattarawut Charatpangoon, Umberto Pensato, Jianhai Zhang, Kazbek Barakhanov, Chitapa Kaveeta, Koji Tanaka, Fouzi Bala, Cody Doolan, Tolulope Sajobi, Brian Buck, Luciana Catanese, Aleksander Tkach, Richard H. Swartz, Nishita Singh, Mohammed A. Almekhlafi, Bijoy K. Menon, Aravind Ganesh

## Abstract

**Background:** Net Water Uptake (NWU) is a non-contrast CT (NCCT) biomarker of early cerebral edema in ischemic stroke, calculated from attenuation differences between ischemic and contralateral non-ischemic brain regions. Manual NWU quantification is labor-intensive and prone to inter-operator variability, limiting clinical uptake and research scalability. We developed and internally validated a fully automated NWU evaluation pipeline.

**Methods:** We analyzed 24-hour follow-up NCCT scans from the AcT (Alteplase compared to Tenecteplase) trial. Infarcts were automatically obtained by segmentation framework based on a synchronous image-label diffusion probability model. The images and extracted infarcts were registered to the standard MNI152 space, allowing us to mirror the infarct onto the contralateral hemisphere symmetrically, regardless of size or tilt angle. Subsequently, the mirrored region was inversely transformed to return to its original space. Voxels outside the range of 20-80 Hounsfield Units (HU) were excluded to remove non-parenchymal tissue. Automated NWU was computed as the percentage difference in mean HU between infarct and mirrored contralateral regions. The agreement with manually determined NWU was evaluated using Pearson correlation, mean absolute error (MAE), and Bland-Altman analysis.

**Results:** Of 1,327 patients in the trial, 298 (22.5%) met predefined imaging-quality criteria for the manual validation analysis, including well-aligned raw NCCT scans in the axial plane and clear parenchymal infarct segmentations. Automated 24-hour NWU showed excellent agreement with manual measurements (r = 0.99). Mean absolute error was 0.18% (95% CI: 0.01-0.46). Bland-Altman analysis demonstrated minimal bias (0.09%) and satisfactory limits of agreement (-4.05% to +4.24%). Ninety-nine percent of cases fell within ±5% of the manually determined value.

**Conclusions:** Our automated mirrored segmentation pipeline enables accurate and reproducible NWU quantification from routine 24-hour NCCT scans, matching expert manual measurements with minimal bias.

## INTRODUCTION

Acute ischemic stroke is a leading cause of death and disability worldwide (1). Brain edema is a central component of ischemic stroke pathophysiology, reflecting and contributing to ischemic tissue injury and poor functional outcome. Hence, early and reliable quantification of ischemia-associated edema is important for prognostication and treatment selection (2,3).

Net water uptake (NWU) derived from non-contrast CT (NCCT) has emerged as a quantitative marker of early ischemic edema (4-6). NWU reflects the relative reduction in tissue CT density in the ischemic region compared with the contralateral normal brain and provides an estimate of the relative increase in water content within the ischemic core (4,5,7). Higher NWU at baseline, indicating more cerebral edema, has been associated with an increased risk of malignant edema, hemorrhagic transformation, and poor functional outcomes despite successful reperfusion (3,5).

Despite this growing evidence, NWU has not been adopted in routine clinical care (3). The main limitation is practical (5). Conventional NWU measurement requires precise manual segmentation of the infarct region on NCCT, construction of a geometrically matched contralateral region of interest (ROI) by manually mirroring the lesion, and voxel-wise CT densitometry within a fixed tissue attenuation window (3,4,5,8). This manual workflow is labor intensive, sensitive to rater expertise – particularly when scans are tilted and corresponding regions do not line up as expected – and difficult to scale across large multicenter cohorts or integrate into real-time decision support (3,9,10).

Recent work has explored partial automation of the NWU pipeline, including automated lesion segmentation and semi-automated density extraction (3,6,10,11). However, most studies have been limited by modest sample sizes, dependence on single-center protocols, or incomplete validation against high-quality manual reference measurements (3,10,11). In particular, accurate contralateral mirroring is technically challenging: simple midline flipping ignores anatomical asymmetries, ventricular shift, and tilt, all of which can introduce systematic bias into NWU estimates (5,10). Furthermore, accurate automated NWU quantification requires accurate infarct segmentation; many prior studies relied on manual infarct segmentation followed by NWU calculation, thereby introducing the bottleneck of manual segmentation (3).

Large randomized controlled trials with imaging acquired in routine care, such as the Alteplase compared to Tenecteplase (AcT) trial comparing Tenecteplase with Alteplase for intravenous thrombolysis, provide an opportunity to rigorously validate automated NWU in a real-world, multicenter setting (12). In particular, follow-up NCCT scans, often acquired in routine practice following thrombolysis, capture the established infarct, in which lesion boundaries are more clearly seen for manual segmentation by expert raters, while edema remains clinically relevant for NWU calculation.

In this study, we validated a fully automated method of NWU quantification using follow-up NCCT scans from the AcT trial (12). Our primary objective was to determine the level of agreement between automated and manual NWU in a rigorously curated dataset. Our secondary objective was to demonstrate that this automated pipeline can be feasibly applied to a large multicenter dataset, providing a practical framework for integrating quantitative edema metrics into future clinical research and into clinical decision-making.

## METHODS

### Study Design and Ethical Approval

This study used anonymized imaging from the AcT randomized controlled trial, a multicenter Canadian study comparing intravenous thrombolysis with Tenecteplase versus alteplase for acute ischemic stroke treatment (12). The trial was conducted at 22 primary and comprehensive stroke centers across Canada. Ethics approval was obtained from the relevant ethics committees at all participating centers (in Calgary, the lead site, this was the Conjoint Health Research Ethics Board at the University of Calgary, REB18-1101). Patients or their authorized representatives provided written informed consent. Further details of the protocol are published elsewhere (13).

### Data Availability

Deidentified spreadsheet data from the AcT trial may be shared on reasonable request from any qualified investigator following review and approval by the AcT executive committee. A request for access to the data can be made by sending an email together with a research plan to the corresponding author.

### Data Collection and Imaging Protocols

Imaging acquired as part of routine care in all enrolled patients in the AcT trial was analyzed in the imaging core lab at the University of Calgary, producing a heterogeneous NCCT dataset with variation in slice thickness (5 mm, 10 mm), scanner models, head orientation and tilts (4, 12). Follow-up NCCT was obtained according to usual post-thrombolysis practice, typically around 24 hours after treatment. For our analysis, we included the AcT trial follow-up scans available in this early post-treatment window (approximately 24-36 hours) (14). These characteristics provided a large, clinically realistic dataset for developing and testing automated NWU computation.

### Manual workflow for lesion segmentation and NWU evaluation

Manual infarct masks were created on follow-up NCCT by fellowship-trained stroke imaging readers with 3-5 years of experience (UP, KB, CK, KT, FB) using ITK SNAP, a validated platform for anatomically guided 3D segmentation (15). Readers segmented the visible infarct core slice by slice and excluded chronic hypodensity, calcification, ventricular spaces or cerebrospinal fluid, and non-ischemic low attenuation tissue.

These manual masks served as the ground-truth dataset for manual NWU calculation. Each infarct mask was mirrored to the opposite hemisphere using a custom geometric mirroring script. Because purely geometric flips do not account for midline shift, sulcal asymmetry, or slice plane tilt, we manually adjusted each mirrored ROI across axial, sagittal, and coronal planes using 3D Slicer, a validated platform for anatomically guided 3D segmentation to ensure anatomically correct correspondence (16).

Cases were ranked by segmentation quality. Cases selected for inclusion in the validation dataset for direct comparison with our automated pipeline had to meet the following criteria: (1) high quality anatomic correspondence defined as accurate midline symmetry with minimal head rotation, (2) precise alignment between infarct and mirrored contralateral anatomy, (3) absence of prior lesions or artifacts in the reference hemisphere, (4) sharply defined infarct boundaries, and (5) no confounding technical artifacts. This ensured that we could be confident about the “ground truth” manually determined NWU.

### Automated NWU evaluation pipeline

We developed a fully automated pipeline that integrates a coarse-to-fine U-Net architecture for infarct segmentation, an approach widely applied in stroke imaging for high-resolution lesion detection, combined with atlas-based nonlinear registration to generate anatomically matched contralateral ROIs using the International Consortium for Brain Mapping (ICBM) symmetric template and the Advanced Normalization Tools (ANTs) registration framework (17,18,19).

The automated infarct segmentation model was developed by collaborators using a hybrid framework that integrates a coarse-to-fine 3D U-Net architecture with a synchronous image-label diffusion probability model for stroke lesion segmentation on non-contrast CT (20,21). This architecture improves over classical U-Net by incorporating sequential refinement stages first predicting the lesion at low resolution, then progressively integrating higher and higher resolution features (20-22). In parallel, the diffusion-based component models the joint evolution of image and label distributions, improving robustness to noise and enhancing lesion boundary delineation in heterogeneous CT data. Skip connections preserved spatial detail, while multi scale feature extraction improved robustness to heterogeneous CT quality (20-22). Training used 5-fold cross validation, with image preprocessing steps including brain extraction, intensity normalization (data whitening), and separation into training/test folds (21,23). This design allowed the model to learn global infarct geometry and fine anatomical structure. The model was trained on CT scans with manually annotated infarcts in the PRoveIT (Measuring Collaterals With Multi-Phase CT Angiography in Patients With Ischemic Stroke) study (24).

To standardize contralateral ROI generation and eliminate subjectivity, we implemented an atlas-based mirroring system using the ICBM 2009a symmetric atlas and ANTs (Advanced Normalization Tools) for deformable registration. The workflow included: (1) Affine registration of each NCCT to the atlas to correct global orientation, (2) Symmetric Normalization (SyN) nonlinear registration to align sulcal and ventricular anatomy, (3) Transformation of the infarct mask into atlas space, (4) Mirroring across the atlas midsagittal plane using Python, and (5) inverse transformation of the mirrored ROI back into each patient’s native space.

This strategy provides anatomically consistent mirroring, avoids midline flip artifacts, and handles variations in tilt and brain asymmetry more reliably than geometric flipping.

Voxel-wise HU values were extracted from the infarct ROI (HU_ischemic), representing the ischemic tissue, and mirrored ROI (HU_contralateral), representing presumed normal brain parenchyma, restricted to 20-80 HU to exclude CSF, bone, calcification, and partial volume boundary voxels (4,5,25). The mean HU value was calculated separately for the infarct ROI and the mirrored contralateral ROI. Net water uptake (NWU) was then computed using the standard formula based on the mean HU difference between these two regions (4,5):

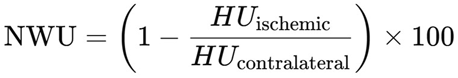

Voxel sizes in the dataset ranged from approximately 0.075 mm³ to 0.4 mm³, depending on scanner and slice thickness (27).

The same formula and HU constraints were applied for both manual and automated NWU to ensure methodological comparability.

After manual agreement validation in the 298-patient AcT validation subset, the automated NWU pipeline was applied to the broader AcT imaging cohort with visible infarcts. Automated outputs were reviewed by core laboratory imaging members (SS, KB) and categorized as acceptable or unacceptable based on segmentation accuracy, contralateral mirroring reliability, and NWU plausibility (Figure 2). Disagreements or uncertain cases were adjudicated by a senior stroke neurologist (AG). This large-scale deployment assessed the generalizability of the automated approach across diverse anatomical and scanner characteristics.

**Figure 1.**
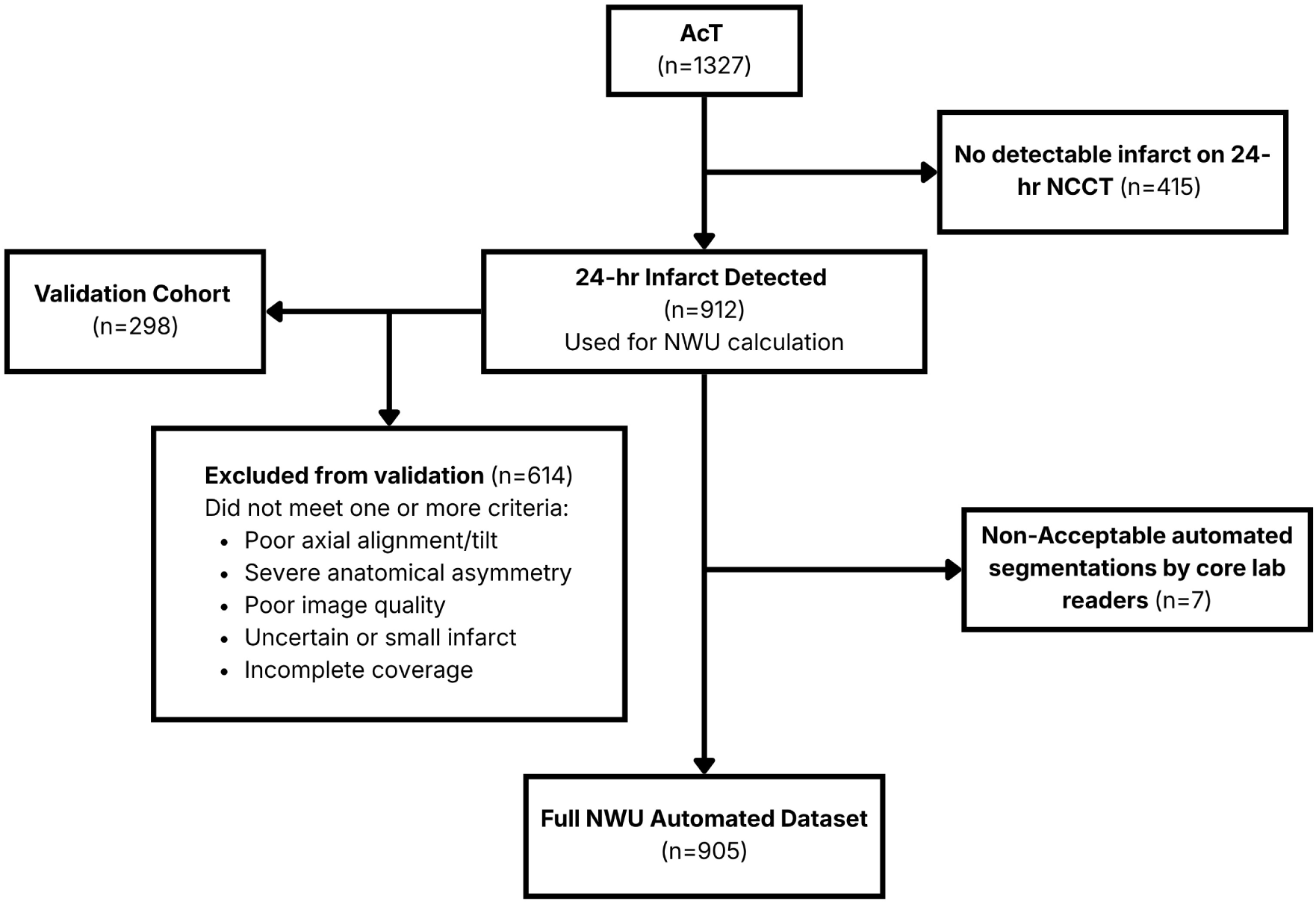
Study Flowchart

**Figure 2.**
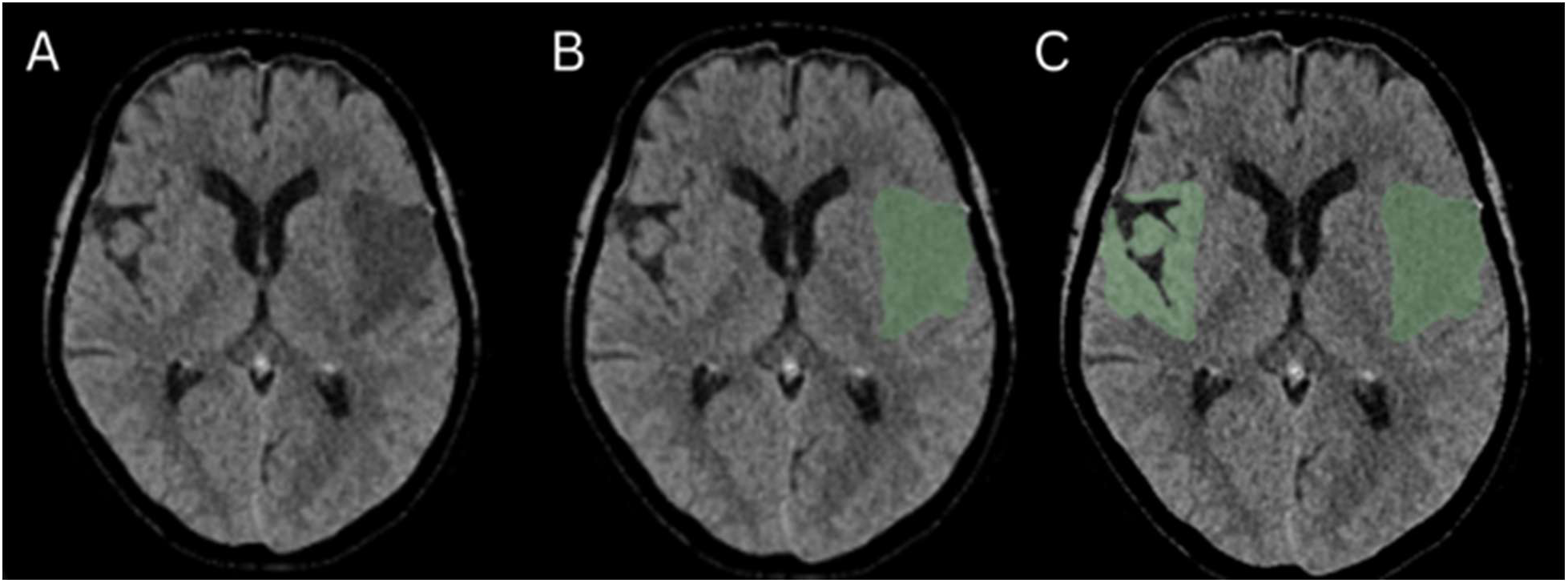
Exemplar patient with acute left middle cerebral artery infarct. A: Raw axial CT slice. B: Automated infarct segmentation (green overlay). C: Infarct segmentation with mirrored contralateral region (green overlay on the radiological right side). Calculated net water uptake (NWU) = 20.13%.

### Statistical Analysis

Agreement between manual NWU and automated NWU was assessed using Pearson correlation coefficient (r), mean absolute error (MAE) with 95% confidence intervals, and Bland-Altman analysis to examine bias and limits of agreement (29).

All analyses were conducted in R (28). Manual and automated HU distributions were also compared using density and histogram analyses to assess systematic differences in ROI sampling. For these distribution analyses, a predefined absolute difference threshold of 5% between automated and manual NWU measurements was arbitrarily used to classify the segmentation error as clinically acceptable. This threshold was selected as a conservative criterion for agreement and is stricter than the ±11.3% limits reported in prior NWU validation studies (3).

## RESULTS

### Patient characteristics

Of 1,577 patients enrolled in the AcT trial, 1,327 had follow-up imaging available for core-lab review. Among these, 912 (68.7%) patients had visible infarcts on follow-up NCCT and were eligible for automated lesion segmentation and NWU calculation. Of these 912 cases, 298 (33%) had clearly delineated infarcts, adequate axial alignment, and reliable contralateral anatomy and were selected for the curated manual-agreement validation subset. After automated processing and core-lab review, 905 of 912 cases were considered acceptable for downstream NWU analysis.

Baseline demographics, treatment workflow variables, and follow-up imaging characteristics for the accepted automated cohort are summarized in Table 1. Characteristics of the 298-patient manual-agreement validation subset are provided in Supplementary Table 1.

**Table 1.** Patient characteristics of the overall study cohort.

|  | **Total Sample**<br>**(n=905)** |
| --- | --- |
| **Demographics** |  |
| Age (years) (median, IQR) | 74 (64-84) |
| Female sex (n, %) | 358 (39.6%) |
| **Baseline features** |  |
| Baseline NIHSS (median, IQR) | 12 (6-18) |
| Baseline ASPECTS (median, IQR) | 9 (8-10) |
| Good collateral status (n, %) | 59 (6.5%) |
| Large vessel occlusion (n, %) | 524 (57.9%) |
| No visible occlusion (n,%) | 166 (18.3%) |
| **Treatment Workflow** |  |
| Tenecteplase use (n, %) | 415 (45.9%) |
| Onset to needle time (minutes) (median, IQR) | 131 (95-190) |
| Onset to arterial puncture time (minutes) (median, IQR) | 160 (126-228) |
| Endovascular thrombectomy use (n, %) | 433 (47.8%) |
| Successful reperfusion (eTICI score $\geq 2b$) on final angiogram run | 213 (23.5%) |
| **Follow-up data** |  |
| 24-hour CT infarct volume (mL) (median, IQR) | 8.7 (1.3-35.9) |
| 24-hour CT infarct density (HU) (median, IQR) | 26.8 (24.0-30.4) |
| 24-hour CT standardized infarct density (sHU) (median, IQR) | 5.6 (4.9-6.8) |

NIHSS National Institute of Health Stroke Scale. ASPECTS: Alberta Stroke Program Early CT Score. eTICI: expanded Thrombolysis in Cerebral Infarction. mL: millilitres. sHU: standardized Hounsfield Unit. IQR: interquartile range.
Large vessel occlusion was defined by baseline CT angiography as occlusions of the intracranial internal carotid artery, first segment (M1) or proximal and dominant second segment (M2) of the middle cerebral artery, or basilar artery.

### Manual-agreement validation in the curated AcT subset

Across the 298 high-quality cases, the mean absolute error (MAE) between automated and manual NWU was 0.18% (95% CI, 0.01-0.46), with a standard error (SE) of 0.12%. The Pearson correlation coefficient demonstrated a strong linear relationship between manual and automated NWU values (r = 0.99, p = 5.36×10⁻243). The relationship is illustrated in Figure 3 (Manual vs Automated NWU scatter plot).

**Figure 3.**
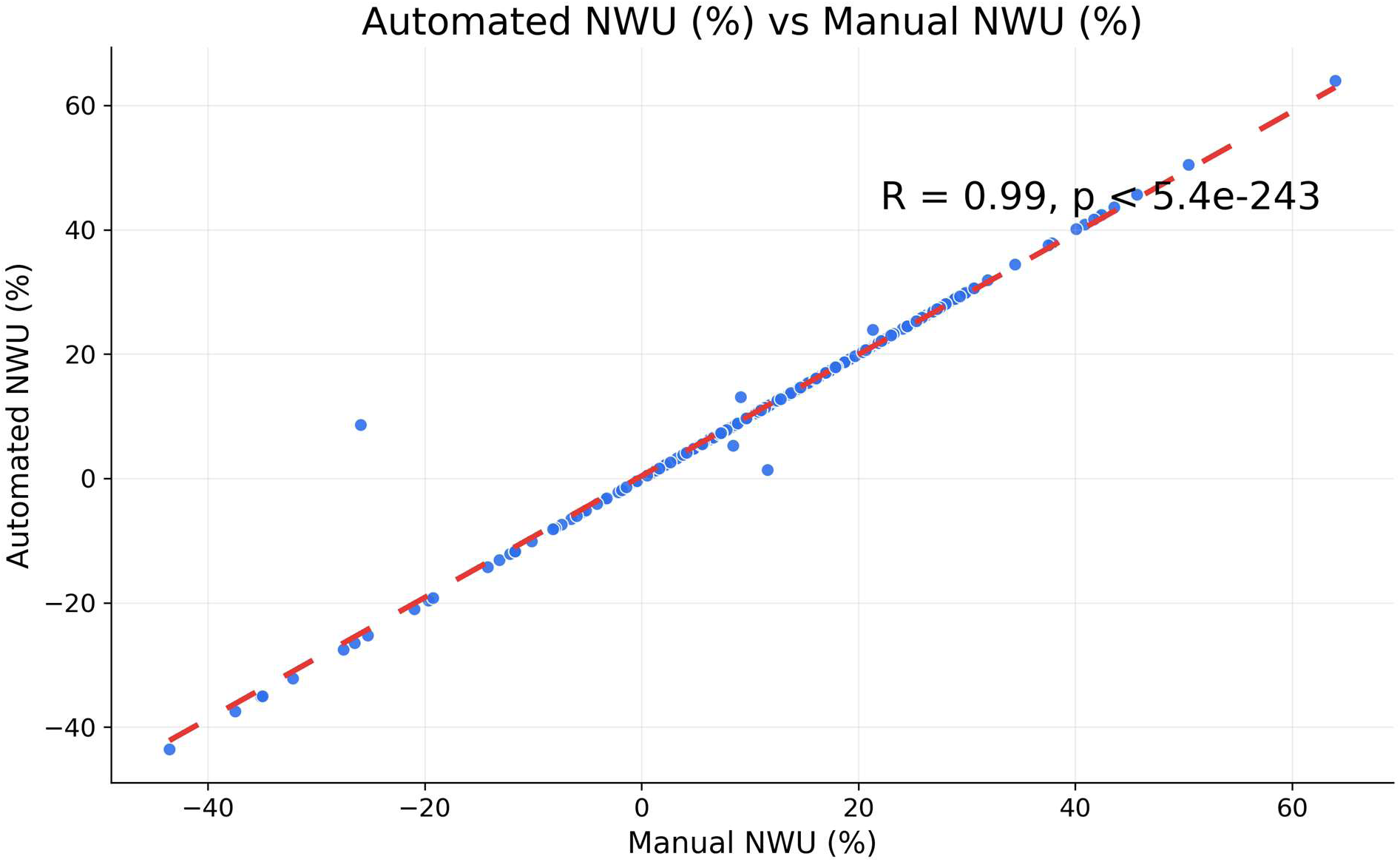
Scatter plot showing correlation between automated-predicted NWU and manually measured NWU across 298 cases. R = 0.99, p = 5.4e-243.

A Bland-Altman plot was constructed to assess systematic bias (Figure 4). The mean bias was 0.09%, indicating that the automated approach produced slightly higher NWU estimates on average. The limits of agreement ranged from -4.05% to +4.24%, consistent with ±1.96 SD (95% CI) and within clinically acceptable tolerance for CT densitometry measures.

**Figure 4.**
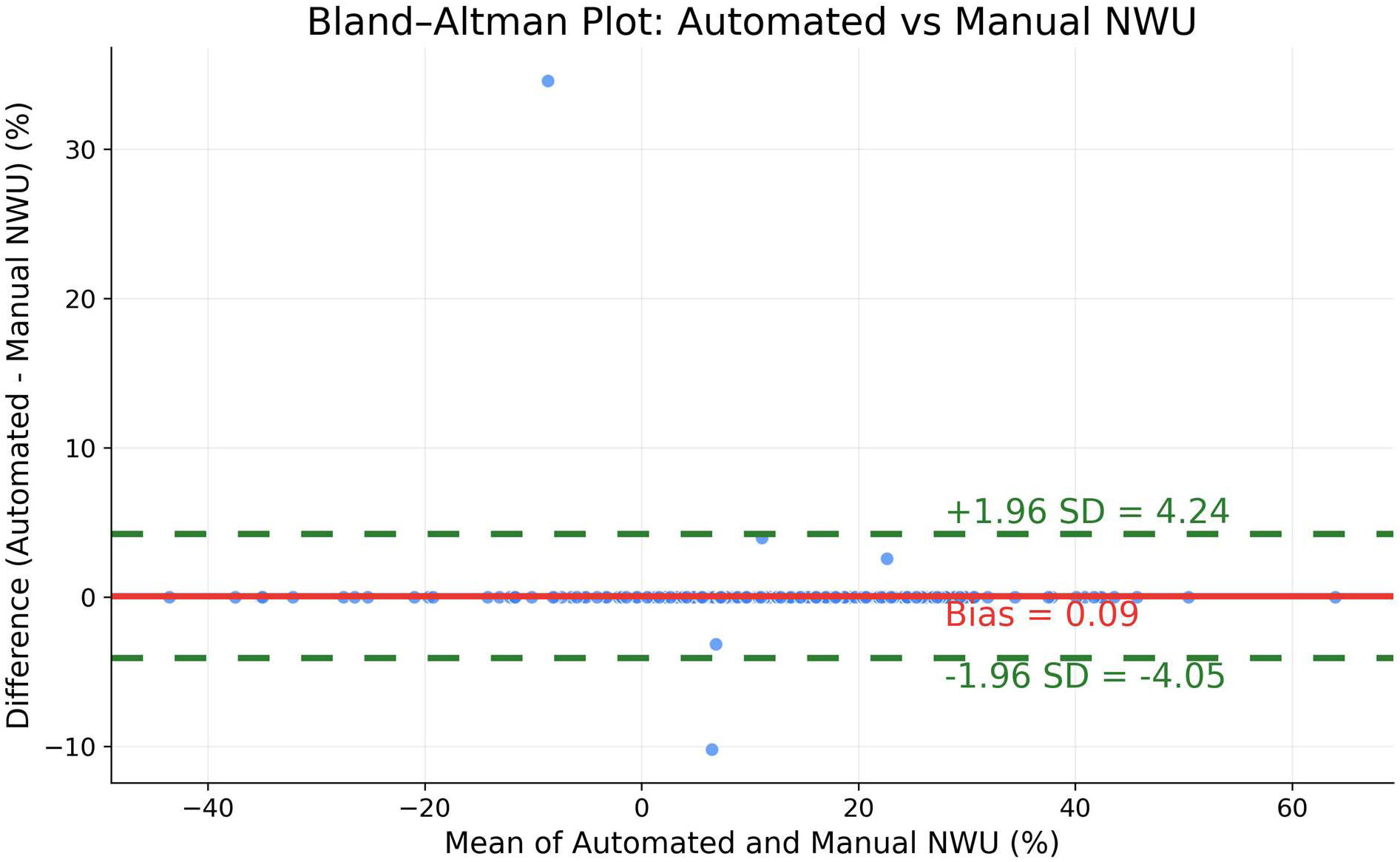
Bland-Altman analysis of automated versus manual NWU. The mean bias between methods was 0.09% (center line). Limits of agreement (±1.96 SD) ranged from –4.05% to +4.24% (green dashed lines).

Error distribution was evaluated using density and histogram analyses (Figure S1 and S2). Absolute differences between manual and automated NWU were right skewed, with most values clustering near the 0.18% error range.

A predefined 5% absolute difference threshold was applied to classify clinical acceptability. 296 of 298 cases (99.33%) fell below the 5% threshold, whereas just 2 cases (0.67%) exceeded the threshold.

### Application to the full cohort

After validation, the model was deployed across all 912 patients in the ischemic stroke dataset. Overall, a total of 905 cases (99.23%, 95% CI 98.42%-99.63%) were considered acceptable by the core lab readers for downstream NWU analysis as shown in Figure 1. Only 7 cases (0.77%) exhibited major errors, including mismatched anatomy, excessive overlap with non-parenchymal structures, or marked registration distortion. The median age was 74 years (IQR=64-84); 358 (51.4%) patients were female; the median baseline NIHSS was 12 (IQR=6-18); and 524 (75.9%) had a large-vessel occlusion. The median 24-hour CT infarct volume was 8.7 mL (IQR=1.3-35.9), the median 24-hour CT standardized infarct density was 26.8 HU (IQR=24.0-30.4), and the median 24-hour CT standardized infarct density was 5.6 standardized HU (IQR=4.9-6.8). Automated NWU values ranged from 0.15% to 48.47% for positive measurements. A small subset of cases (n=81) exhibited negative NWU values, extending down to -66.63%, representing the full observed measurement range. Of these, 41 (50.6%) demonstrated hemorrhagic transformation on follow-up imaging. Overall, the NWU distribution showed a mean of 11.57%; median NWU at 12.86% (IQR: 7.13% - 18.94%); SD (12.44%) (95% CI for mean NWU: 10.75% - 12.38%) (Figure S3).

## DISCUSSION

In this multicenter analysis of follow-up NCCT imaging from the AcT trial, we found that a fully automated pipeline for quantifying 24-hour net water uptake (NWU) closely approximated expert manual assessment (12). Three main findings emerged. First, automated NWU demonstrated excellent agreement with manual ground-truth values in the curated validation subset, with a correlation coefficient of r = 0.99 and a mean absolute error of approximately 0.18 percentage points. Second, Bland–Altman analysis showed minimal systematic bias, supporting the reliability of automated HU-based edema quantification. Third, when extended to a large, heterogeneous cohort of 912 patients, the automated workflow maintained acceptable segmentation, anatomical mirroring, and NWU plausibility in more than 99% of cases. Together, these results indicate that fully automated 24-hour NWU computation is technically feasible, reproducible, and scalable for use in large imaging datasets.

The present study confirms that a fully automated approach can reproduce these quantitative measures with clinically acceptable precision. That the mean bias was 0.09% indicating only a slight overestimation by the automated method suggests that atlas based mirroring and standardized HU sampling preserve the fidelity of NWU, even in the presence of anatomical asymmetry or variable slice orientation.

The distribution of errors provides additional insight. Most absolute differences were clustered near 0.18%, and more than 99% of cases fell within a conservative 5% threshold. These limits of agreement were narrower than the ±11.3% reported in a prior multicentre imaging study of automated NWU measurement, although direct comparison is limited because prior work assessed baseline NWU, where ischemic hypoattenuation may be subtler and more difficult to segment than on 24-hour follow-up imaging (3, 5). Together, these findings support the robustness of the pipeline for follow-up NCCT-based NWU quantification across diverse imaging conditions. Outlier cases, which exceeded the 5% threshold, appeared to have pronounced anatomical distortion, excessive axial tilts, or complex lesion morphology. These scenarios underscore the need to retain human oversight of automated outputs in specific cases, particularly when quantitative CT metrics are used to inform acute clinical decisions.

Accurate NWU quantification depends critically on the integrity of both infarct segmentation and contralateral region selection. Variability in CT slice thickness and axial tilt poses inherent challenges for quantitative imaging on routine clinical scans. In our study, these issues were prominent in the manual workflow, requiring extensive adjustment of mirrored ROIs to correct for asymmetry, ventricular displacement, and partial volume artefacts. By contrast, the automated pipeline’s use of atlas-based nonlinear registration offered more consistent anatomical correspondence and reduced the dependency on slice-specific corrections. The atlas-based approach also benefits from predictable behavior across heterogeneous datasets, making it more suitable for multicenter trials than a purely geometric midline flip.

The automated pipeline was built on the same HU thresholds (20-80 HU) and NWU formula used in foundational work by previous studies, restricting HU values to this range minimizes contamination by CSF, necrotic tissue, hemorrhage, and bone, thereby stabilizing NWU calculations across variable acquisition protocols (4,5,25). The consistency between our NWU distributions and those reported in previous studies, mean values around 10%-15% for moderate to severe edema further supports the biological plausibility of the automated measurements (3,5).

This study also contributes new evidence regarding the feasibility of deploying automated NWU tools at scale. Among the 912 processed cases, fewer than 1% demonstrated major segmentation or registration failure. Such a low failure rate for a fully automated pipeline is noteworthy given the heterogeneity of the AcT dataset, which included variable scanner models, slice thicknesses, and degrees of head rotation (12).

Although most cases demonstrated positive NWU values consistent with expected ischemic edema patterns, a small number of acceptable cases exhibited negative NWU values. These negative values likely reflect situations in which the ischemic region demonstrated higher attenuation than the contralateral reference tissue. Such findings may occur in the presence of hemorrhagic transformation, localized hyperattenuation, or contrast staining, which can increase attenuation within infarcted tissue on CT, as well as from segmentation-related variability in HU sampling within heterogeneous infarct regions (2,5,8). Importantly, these cases represented a small minority and did not materially affect the overall NWU distribution.

Overall, the NWU distribution was physiologically plausible and aligned with expected edema severity ranges. The automated system therefore shows promise for integration into potentially real-time clinical pipelines. As quantitative stroke imaging continues to evolve, automated NWU may complement perfusion imaging, collateral scoring, and advanced infarct characterization to support treatment selection and prognostication (3,6,30,31).

Several limitations warrant consideration. First, NWU was calculated on follow up NCCT, and hyperdense changes such as contrast retention or microbleeds can raise local HU values (3). These effects may make edema appear smaller than it truly is. Although our HU thresholds mitigate this effect, dual energy CT (DECT) or virtual non contrast reconstructions may offer more robust differentiation of blood, contrast, and tissue, and should be evaluated in future work (32). Second, imaging parameters varied across centers, reflecting the pragmatic nature of the AcT trial (12). While this heterogeneity strengthens external validity, it may introduce variance in HU measurements and segmentation performance. Third, although the validation dataset was curated to ensure high anatomical quality, real-world clinical implementation will require safeguards for detecting and managing problematic cases. Finally, the present study focused on accuracy of NWU calculation rather than outcome prediction; future analyses should examine how automated NWU relates to functional and radiologic endpoints, including malignant edema, reperfusion injury, and hemorrhagic transformation. In parallel, ongoing work from our group will evaluate whether NWU measured at baseline and on follow-up CT adds prognostic value beyond infarct volume and infarct density in outcome modeling across the AcT trial cohort.

## CONCLUSION

Automated quantification of 24-hour net water uptake (NWU) demonstrated excellent agreement with expert manual measurements in this multicenter AcT cohort. The atlas-registered mirrored segmentation pipeline enabled accurate and reproducible NWU estimation from routine non-contrast CT with minimal bias. By reducing manual workload and supporting standardized edema quantification, this approach provides a scalable imaging biomarker for stroke research, future clinical trials, and imaging-based prognostic modeling.

## Non-standard abbreviations and acronyms

NWU: Net Water Uptake
HU: Hounsfield Unit
NCCT: Non Contrast CT

## ACKNOWLEDGEMENTS

None

## SUPPLEMENT

**Supplementary Table 1.** Patient characteristics of the manual validation cohort.

|  | **Total Sample**<br>**(n=298)** |
| --- | --- |
| **Demographics** |  |
| Age (years) (median, IQR) | 73 (62-83) |
| Female sex (n, %) | 115 (38.6%) |
| **Baseline features** |  |
| Baseline NIHSS (median, IQR) | 12 (6-19) |
| Baseline ASPECTS (median, IQR) | 8 (7-10) |
| Good collateral status (n, %) | 13 (4.4%) |
| Large vessel occlusion (n, %) | 181 (60.7%) |
| No visible occlusion (n, %) | 47 (15.8%) |
| **Treatment Workflow** |  |
| Tenecteplase use (n, %) | 132 (44.3%) |
| Onset to needle time (minutes) (median, IQR) | 132 (93-194) |
| Onset to arterial puncture time (minutes) (median, IQR) | 161 (125-236) |
| Endovascular thrombectomy use (n, %) | 134 (45.0%) |
| Successful reperfusion (eTICI score $\geq 2b$) on final angiogram run | 72 (24.2%) |
| **Follow-up data** |  |
| 24-hour CT infarct volume (mL) (median, IQR) | 12.2 (2.3-39.0) |
| 24-hour CT infarct density (HU) (median, IQR) | 27.8 (24.0-31.6) |
| 24-hour CT standardized infarct density (sHU) (median, IQR) | 6.0 (4.9-7.1) |

**Supplementary Figure 1.**
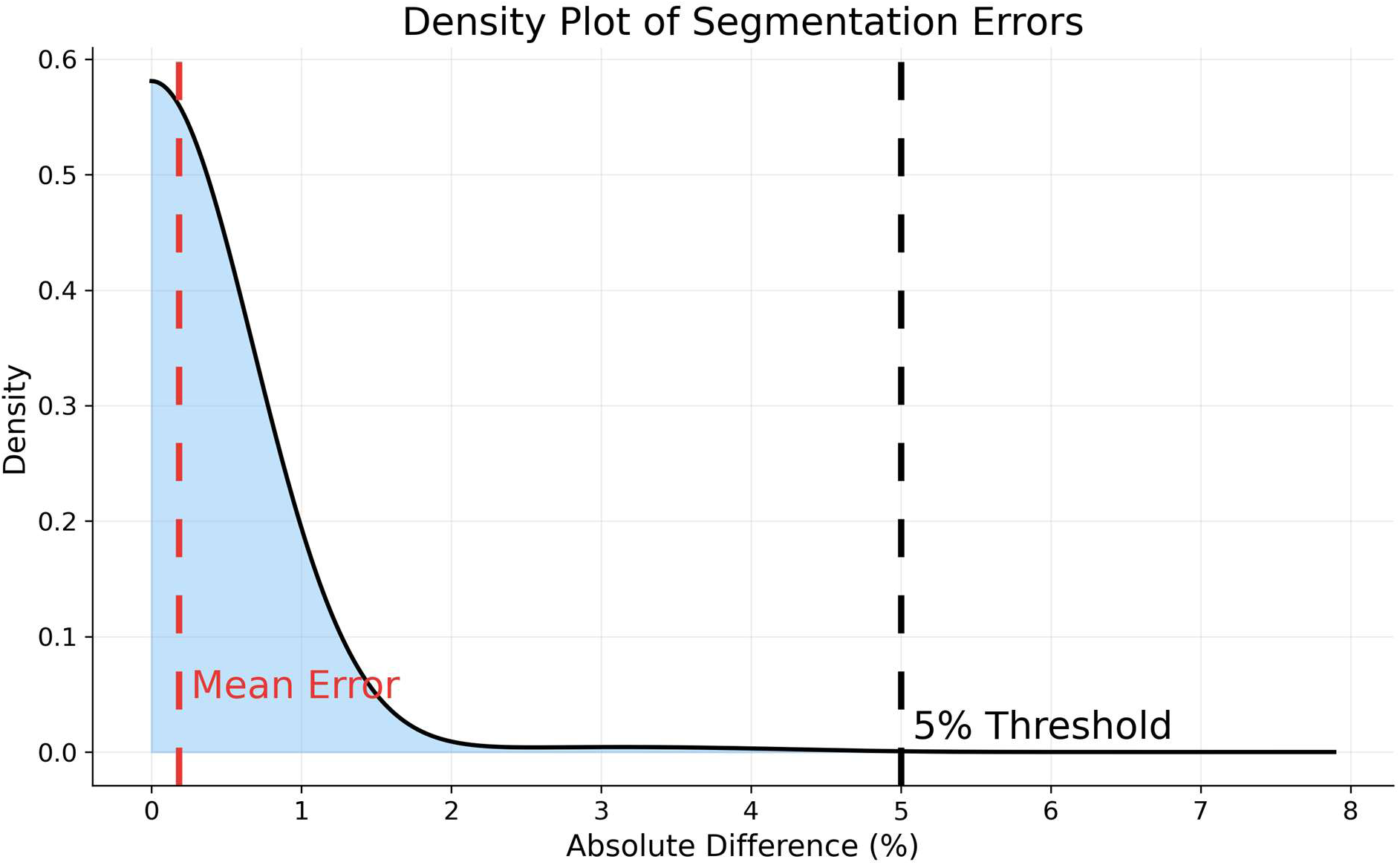
Density plot showing distribution of absolute segmentation error (in %). Mean error ≈ 0.18%, with most values clustered below the 5% clinical threshold.

**Supplementary Figure 2.**
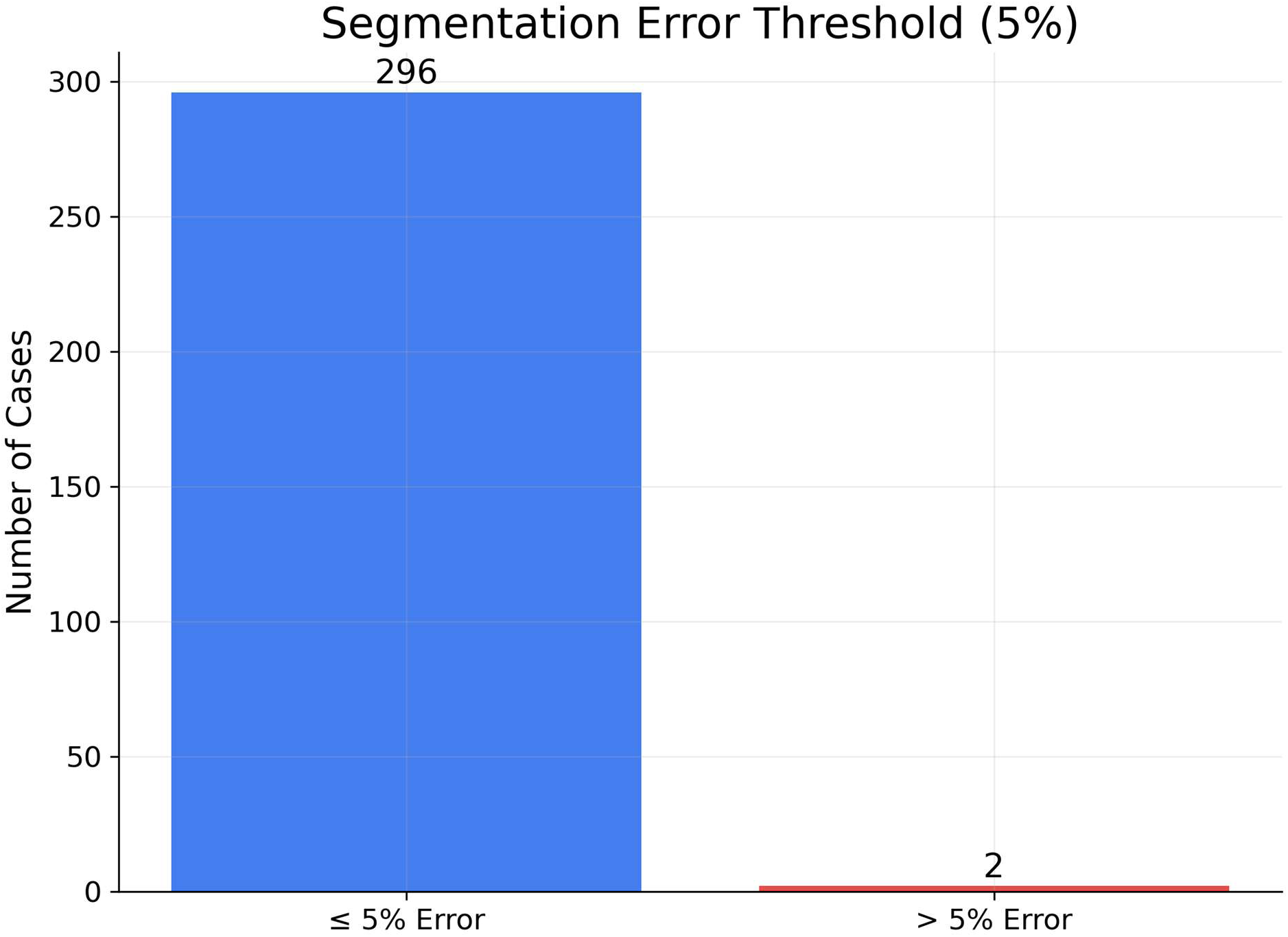
Bar chart of segmentation error categories: Below vs. Above 5% threshold. 296 cases below 5% threshold; 2 cases above.

**Supplementary Figure 3.**
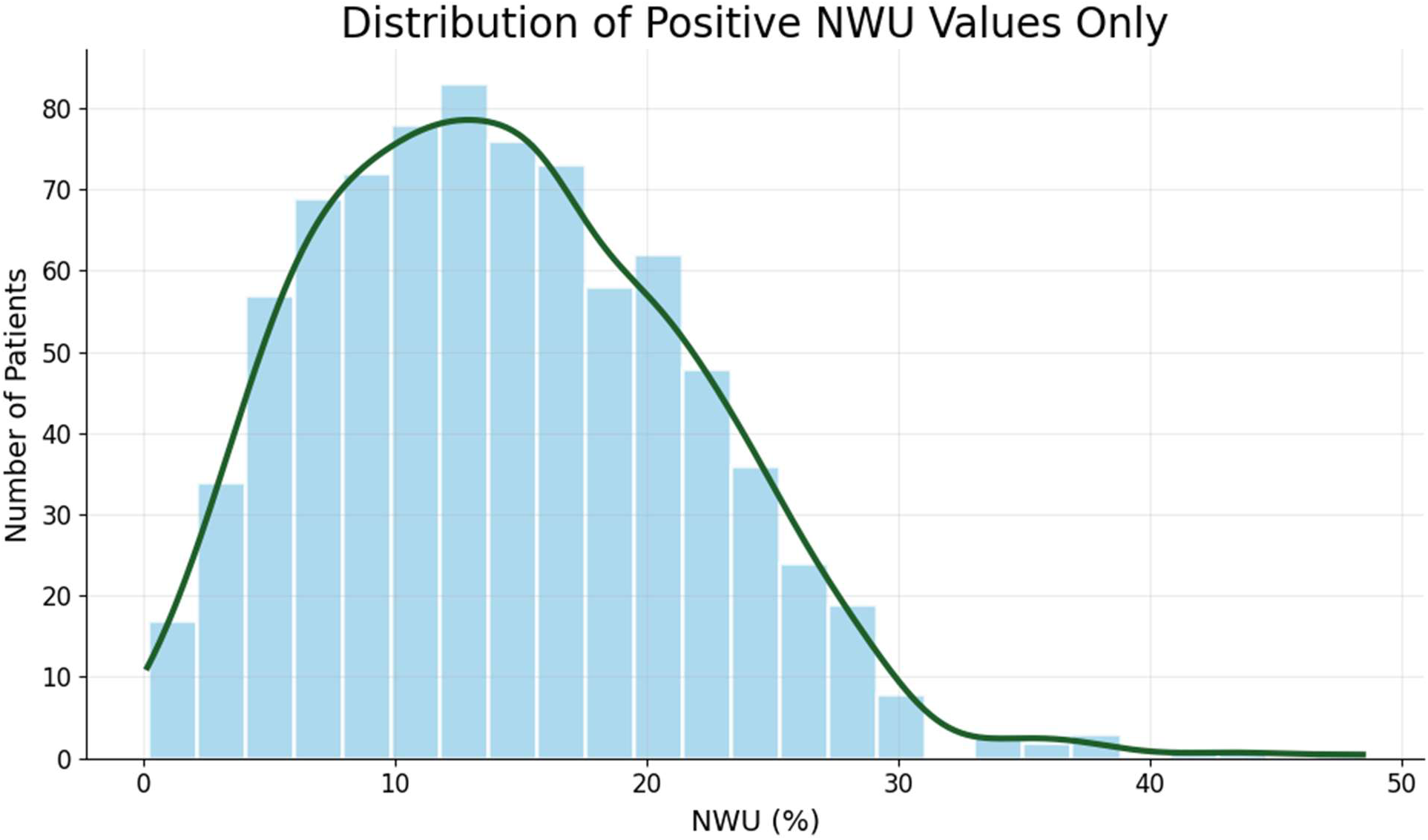
Histogram and density plot showing the distribution of positive net water uptake (NWU) values (n=825) among the 905 automated cases. The distribution demonstrates a right-skewed pattern, with most NWU measurements concentrated between 5-20%, consistent with expected edema severity ranges on follow-up NCCT.

